# Pro-ictal EEG scheduling improves the yield of epilepsy monitoring: Validating the use of multiday seizure cycles to optimize video-EEG timing

**DOI:** 10.1101/2023.11.27.23299049

**Authors:** Jodie Naim-Feil, Rachel E. Stirling, Philippa J. Karoly, Daniel Payne, Nicholas Winterling, Dominique Eden, Mark J. Cook, David B. Grayden, Matias Maturana, Dean R. Freestone, Ewan S. Nurse

**Author notes:** **Correspondence:** Dr Jodie Naim-Feil.

## Abstract

A significant challenge of video-electroencephalography (vEEG) in epilepsy diagnosis is timing monitoring sessions to capture epileptiform activity. Given the significant consequences of misdiagnosis or delayed diagnosis, new techniques to improve diagnostic yield of vEEG are needed. In this study, we introduce and validate “pro-ictal EEG scheduling”, a method to schedule vEEG monitoring to coincide with periods of heightened seizure probability as a low-risk approach to enhance the diagnostic yield. A database of long-term ambulatory vEEG monitoring sessions (*n*=5038) of adults and children was examined. Data from linked electronic seizure diaries were extracted (minimum 10 self-reported events over 12-months) to generate cycle-based estimates of seizure risk. VEEG monitoring sessions coinciding with periods of estimated high-risk were allocated to the high-risk group (adults *n*=305, children *n*=82) and compared to remaining studies (baseline: adults *n*=3586, children *n*=1065). Test of Proportions and Risk-Ratios (RR) were used to index differences in proportions and likelihood of capturing outcome measures (abnormal report, confirmed seizure and diary event) during monitoring. The impact of clinical and demographic factors (sex, epilepsy-type, medication) was also explored. During vEEG monitoring, the high-risk group was 25% more likely to have an abnormal vEEG report (190/305:62.3% vs 1790/3586:49.9%, RR=1.25, 95% CI[1.137:1.370], *p*<0.001), 63% more likely to present with a confirmed seizure (56/305:18.4% vs 424/3586:11.3%, RR=1.63, 95% CI[1.265:2.101], *p*<0.001) and 42% more likely to report an event (153/305:50.2% vs 1267/3586:35.3%, RR=1.420, 95% CI[1.259:1.602], *p*<0.001). In children, the high-risk group was 93% more likely to have a confirmed seizure (21/82:25.6% vs 141/1065:13.2%, RR=1.93, 95% CI[1.297:2.885], *p=*0.002). Similar effects were observed across clinical and demographic features. This study provides the first large-scale validation of pro-ictal EEG scheduling in improving the yield of vEEG. This innovative approach offers a pragmatic and low-risk strategy to enhance the diagnostic capabilities of vEEG monitoring, significantly impacting epilepsy management.

## Introduction

Video-electroencephalography (vEEG) is a well-established diagnostic tool for identifying epilepsy syndromes and characterising the seizure onset zone for pre-surgical planning. While there is broad consensus of the diagnostic utility of vEEG in capturing both epileptic and non-epileptic seizures ^1^, data from large-scale retrospective studies observed that, in patients previously diagnosed with epilepsy, vEEG was unable to capture seizures or typical episodes across both adult^1,2^ and paediatric cohorts^3^ in up to 30% of cases. Misdiagnosis, diagnostic delay and inconclusive results can have a diverse range of adverse effects. This includes sub-optimal clinical management (and unsuitable medication regimen) and potential life-threatening injury from undiagnosed epilepsy, as well as considerable psychological and social impact (including potential employment and driving restrictions) and further delays in identifying the source of potential nonepileptic events (i.e., cardiac, PNES and psychiatric causes) ^4–7^. Moreover, if the diagnosis is not conclusive, there is a significant personal and economic impact (including costs associated with hospital infrastructure and personnel) of repeated hospital admissions and assessments ^7,8^. All these factors are a significant burden on individuals living with epilepsy and the supporting health care systems.

Given the substantial consequences of misdiagnoses, delayed diagnosis, and repeated admissions in epilepsy, developing techniques to improve the diagnostic yield of vEEG is crucial to mitigating these risks. Various methods are deployed to increase the yield of vEEG for the diagnosis of epilepsy and presurgical planning, such as medication withdrawal ^2^, hyperventilation ^9^, photic stimulation ^10^ and sleep deprivation ^11^. While these seizure-provoking techniques can be effective, they require highly specialized, supervised hospital settings to monitor seizure activity due to the increased risk of seizure clusters and potential for status epilepticus ^12^. Here, we introduce and validate the concept of “pro-ictal EEG scheduling”, or timing EEG monitoring to coincide with periods of heightened seizure risk, as a low-risk strategy to increase the clinical yield of vEEG.

This study aims to validate a potential approach for pro-ictal EEG scheduling based on multiday seizure cycles^13–16^. Advances in long-term electroencephalography monitoring ^17^ have provided critical insights into the multiday temporal dynamics underlying seizure occurrence ^13–16^, presenting compelling electrographic evidence that electrographic seizures^15–18^ and interictal epileptiform discharges ^14,19,20^ follow patient-specific cyclic patterns across both circadian and multiday timescales. A significant benefit in the discovery of the cyclic nature of these seizure patterns is the potential to project multiday cycles forward ^18,21^ to estimate future periods of heightened seizure risk (i.e., the pro-ictal state) across a range of timescales (days, weeks, months) ^15,22–24^. Recently, researchers demonstrated that multiday seizure cycles can also be extracted from seizure diaries ^25–27^ with comparable cycles to electrographic seizure patterns observed in intracranial recordings ^18,20^, and can be projected forward to estimate future periods of heightened seizure risk ^13,20,28^. These significant advances in seizure forecasting hold valuable clinical potential for improving the diagnostic effectiveness of vEEG by timing vEEG monitoring to coincide with projected pro-ictal periods. Thus far, this novel approach has been assessed in a single retrospective cohort study involving 48 participants ^28^, providing the first line of evidence that vEEG monitoring conducted during the pro-ictal state aligns with a higher rate of conclusive diagnostic outcomes and heightened epileptiform activity^28^.

In this study, we introduce the concept of pro-ictal EEG scheduling as a promising low-risk framework to enhance the diagnostic efficacy of vEEG, potentially reducing the risk of misdiagnosis and diagnostic delays. To validate this approach, the current study evaluates whether personalized predictions of heightened seizure risk (as derived from electronic seizure diaries) align with increased likelihood of capturing epileptiform activity and reported events during vEEG monitoring. This evaluation is conducted using data extracted from a database of more than 5000 ambulatory long-term vEEG sessions. Subsequently, the model is validated within a paediatric cohort. Following this, the potential impact of clinical and demographic features in modulating the relationship between pro-ictal scheduling and diagnostic outcome measures during vEEG monitoring is investigated.

## Materials and methods

In this large-scale study, a database (Seer Medical Pty Ltd, Australia) of long-term ambulatory vEEG monitoring sessions (*n*=5038) with linked electronic seizure diaries is examined. The following section outlines how multiday forecasts of seizure risk were generated from seizure diaries to examine whether timeframes of forecasted high-risk correspond to vEEG outcomes relative to a baseline comparison. The study pipeline is depicted in Figure 1.

**Figure 1.**
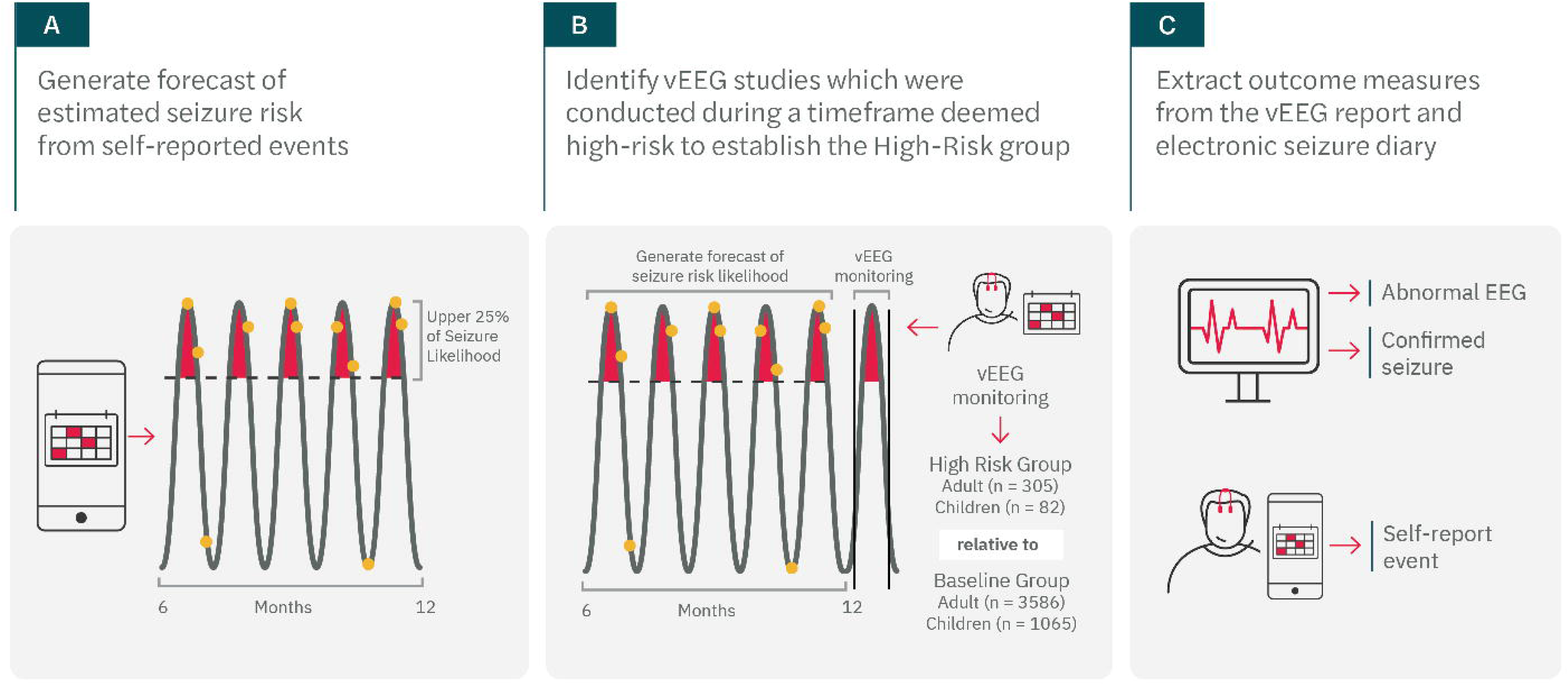
Schematic visualization of the study pipeline. **A.** Data were analysed from the large ambulatory vEEG database of adults and children with a linked electronic seizure diary of any length. For each person, estimated seizure likelihood was calculated based on self-reported events logged into an electronic seizure app and seizure forecasts generated for participants who presented with a minimum of 10 reported events over a 12-month period. **B.** All vEEG studies that were conducted during a period of forecast high-risk (>25% of time spent above the high-risk threshold during vEEG monitoring) were identified and were allocated into a subgroup called the “high-risk group”. All other vEEG studies were considered to be the comparison “baseline group”. **C.** Outcome measures were extracted from the EEG report, which included identification of whether the EEG was normal or abnormal (i.e., epileptic activity identified) and the presence of confirmed electrographic seizures during vEEG monitoring. Participant self-reported diary event events during vEEG monitoring were also extracted. Self-reported seizures are indicated by orange dots and the upper 25% of seizure likelihood is represented by red shading above the dotted black threshold line. The solid oscillating line represents the multiday seizure cycle calculated from the seizure diary prior to the vEEG monitoring period.

### Participants

In this study, vEEG monitoring studies of both adults (≥18 years) and children (<18 at time of monitoring) conducted between 2019-2022, with a linked electronic seizure diary, were included for analysis. For each person, estimated seizure likelihood was calculated based on self-reported events logged into an electronic seizure app prior to the vEEG monitoring sessions. Forecasts detailing the estimated seizure likelihood were generated for participants who presented a minimum of 10 reported events over a 12-month period (further described below in Identifying cyclic patterns of seizure risk). The high-risk threshold was defined as the top 25^th^ percentile of all past likelihoods. All vEEG studies that were conducted during a period of forecast high-risk (>25% of time spent above the high-risk threshold during vEEG monitoring) were included in the group called the “high-risk group”. All other vEEG studies were allocated to the comparison “baseline group”. De-identified data for this study was stored on a cloud platform and derived from 1) the vEEG report (vEEG outcome, whether there was a confirmed electrographic seizure, type of epilepsy including generalized or focal based on neurologist review of epileptiform discharges/spikes and confirmed seizures), 2) registration form (age, sex, seizure frequency), 3) the clinician referral form (medications), and 4) self-reported seizure events logged in a linked electronic seizure diary.

Ethical approval for this study was granted by the St Vincent’s Hospital Melbourne Human Research Ethics Committee (165/19). Participants provided written informed consent for their de-identified data to be used for this study prior to their vEEG monitoring.

#### Video-electroencephalography (vEEG)

The vEEG monitoring sessions were conducted using an ambulatory vEEG system^29^. During vEEG monitoring, patients were instructed to move the mobile camera system with them throughout their home environment (excluding bathroom). No seizure provocation methods, such as medication withdrawal or sleep deprivation, were used during monitoring. Suspected interictal epileptiform events were identified via a machine learning algorithm ^30^. All suspected events were manually reviewed on a cloud platform by a trained neurophysiologist and any abnormalities were noted for the EEG outcome assessment (including focal slowing, polyspikes, spike/sharp/slow-wave complexes, and the presence of rhythmic and periodic activity in the EEG recording). Identification of electrographic seizures was based on visual review of the EEG traces and was confirmed (where possible) by the simultaneous video recording. This report (and the vEEG recording) was also reviewed by an expert neurologist, who provided a final assessment of the presence of any epileptiform activity during vEEG monitoring. Based on this assessment, the neurologist provided a concluding report defining the vEEG outcomes as normal (no evidence of epileptic activity) or abnormal (i.e., identified epileptiform activity and/or confirmed seizures).

#### Seizure diaries

Seizure times were recorded in an electronic seizure diary smartphone app. Recorded information included the date and time of the event, seizure type, symptoms, and duration of event. Date and time of the seizure events were extracted: 1. across the mobile diary from activation up until 1 week prior to the vEEG monitoring (to generate seizure risk forecast) and 2. during the vEEG monitoring session, with logged diary events during monitoring considered a “self-reported event” outcome measure.

#### Identifying cyclic patterns of seizure risk

Seizure cycles estimated from an individual’s self-reported event times were extracted to determine periods of high seizure risk. A probabilistic framework for seizure likelihood was applied, whereby the probability of seizure occurrence with respect to each cycle was calculated based on a circular histogram of reported seizure event times^18^. Briefly, the strongest “fast” (2-7 days) and “slow” (7.5-70 days) cycles were established based on phase locking of self-reported seizure times. Phase locking was quantified using the synchronisation index (SI), as recommended for analysis of seizure cycles^21^. Where possible, seizure cycles were extracted prospectively (*n* = 174, diary events were reported prior to vEEG monitoring), but, if unable to extract a seizure cycle prospectively (i.e., not enough events logged prior to vEEG monitoring), diary events following vEEG monitoring (*n* = 131) were used to establish seizure cycles. Any seizure events logged during or one week before or after vEEG monitoring were not included in the identification of seizure cycles nor determination of high-risk periods. The high-risk threshold was set as the top 25^th^ percentile of all past likelihoods, with all EEG datasets having >25% of time above the high-risk threshold during vEEG monitoring session allocated to the “high-risk group”.

### Statistical analysis

For the primary analysis, the independent variable was risk level (high-risk or baseline), and the dichotomous dependent outcome measures (each assessed separately) were: 1. EEG report outcome (normal/abnormal), 2. presence of confirmed seizure (≥1) during vEEG monitoring, and 3. self-reported diary events logged (≥1) during the vEEG monitoring. Test of Two Proportions (chi-square test for homogeneity) was applied to identify if a difference existed between the binomial proportions (of the outcome measures) between the two risk groups (high-risk and baseline comparison) and whether the difference in proportions was significant (*p* < 0.05). The Relative-Risk Ratio (RR) was also reported to identify the likelihood of an outcome occurring in the high-risk group compared to the probability of it occurring in the baseline comparison group.

For the secondary analysis, the groups were further divided according to 1. sex (female/male), 2. epilepsy type (generalized/focal) and 3. medication (anti-seizure medications reported in clinical referral/no anti-seizure medications reported), and the strength of association of the risk groups in increasing the likelihood of identifying the three diagnostic outcome variables was examined. The primary analysis was conducted across adults and children separately. All secondary analyses were conducted in the adult group only, as the sample size was too small to conduct the secondary analyses in subgroups of the paediatric cohort.

All statistical analysis was conducted using SPSS statistics for Windows (Version 27.0).

## Results

### I. Demographic features

Comparability of the high-risk group (adults: *n*=305, children: *n*=82) to the baseline group (adults: *n*=3586, children: *n*=1065) was assessed by χ²-tests for categorical variables (sex and medication) and independent *t*-tests for continuous variables (age), while study duration (length in days) comparisons were conducted using the Mann-Whitney U test and the independent samples Median test to address violations of normality in the distribution (Table 1). Notably, given the significant differences in mean days of study duration and the percentage medicated with anti-seizure medication across the risk groups, these effects are explored more thoroughly in *Section II. Secondary Analysis*.

**TABLE 1.** Demographic and clinical data for all participants for each risk group divided according to Age group (Adults: ≥18 years and Children: < 18 years).

|  | Adults |  | Children |  |
| --- | --- | --- | --- | --- |
| | High-Risk<br>(*n* = 305)<br>Mean ± SD | Baseline<br>(*n* = 3586)<br>Mean ± SD | High-Risk<br>(*n* = 82)<br>Mean ± SD | Baseline<br>(*n* = 1065)<br>Mean ± SD |
| **Age (years)**<br>*p*-value | 39 ± 15<br>*p* = 0.115 | 40 ± 16 | 12 ± 4<br>*p* = 0.555 | 12 ± 4 |
| **Sex (Female: Male)**<br>*p*-value | 193 : 90<br>*p* = 0.195 | 2204 : 1210 | 55 : 25<br>*p* = 0.116 | 612 : 411 |
| **Study duration (days)\***<br>*p*-value | 6 ± 2<br>Median [Range] = 7 [2:10]<br>*U*: *p* < 0.001, Median: *p* = 0.120 | 6 ± 2<br>Median [Range] = 7 [1:10] | 5 ± 2<br>Median [Range] = 4 [1:10]<br>*U*: *p* = 0.085, Median: *p* = 0.188 | 4 ± 2<br>Median [Range] = 3 [1:10] |
| **% Medicated with ASM**<br>*p*-value | 185 (61%)<br>*p* < 0.001 | 1804 (50%) | 44 (54%)<br>*p* = 0.014 | 424 (40%) |
| **% Number of concurrent ASMs** | 1 ASM: 75/305: 25%<br>2 ASM: 47/305: 15%<br>3 ASM: 40/305: 13%<br>4+ ASM: 23/305: 8% | 1 ASM: 959/3586: 27%<br>2 ASM: 509/3586: 14%<br>3 ASM: 228/3586: 6%<br>4+ ASM: 108/3586: 3% | 1 ASM: 75/82: 29%<br>2 ASM: 47/82: 18%<br>3 ASM: 40/82: 5%<br>4+ ASM: 23/82: 1% | 1 ASM: 959/1065: 22%<br>2 ASM: 509/1065: 12%<br>3 ASM: 228/1065: 4%<br>4+ ASM: 108/1065: 3% |
| **% Common ASMs\*\*** | Carbamazepine (19%)<br>Lamotrigine (30%)<br>Levetiracetam (42%)<br>Topiramate (13%)<br>Valproate (30%) | Carbamazepine (17%)<br>Lamotrigine (31%)<br>Levetiracetam (39%)<br>Topiramate (13%)<br>Valproate (27%) | Carbamazepine (23%)<br>Lamotrigine (20%)<br>Levetiracetam (36%)<br>Topiramate (7%)<br>Valproate (32%) | Carbamazepine (13%)<br>Lamotrigine (26%)<br>Levetiracetam (33%)<br>Topiramate (9%)<br>Valproate (33%) |

\*For study duration (days), both a t-test and an independent Sample Median Test was conducted to compare the 1. Mean and 2. Median between the risk categories. \*\*Patients may be on multiple medications. All medications refer to anti-seizure medications reported on the referral form by the clinician. Only those who recorded Sex as Female/Male are included in the table above. Abbreviations: Anti-Seizure Medication (ASM).

### I. Primary Analysis

#### Adults

3891 adults who completed ambulatory vEEG monitoring were divided into the high-risk forecast (*n*=305) or baseline (*n*=3586) condition (as presented in Figure 2A). Relative to baseline, those in high-risk were 25% more likely to have an abnormal report (190/305:62.3% vs 1790/3586:49.9%, RR=1.25, 95% CI[1.137:1.370], *p*<0.001), 63% more likely to present with a confirmed seizure during vEEG monitoring (56/305:18.4% vs 424/3586:11.3%, RR=1.63, 95% CI[1.265:2.101], *p*<0.001) and 42% more likely to report a diary event during monitoring (153/305:50.2% vs 1267/3586:35.3%, RR=1.420, 95% CI[1.259:1.602], *p*<0.001).

**Figure 2.**
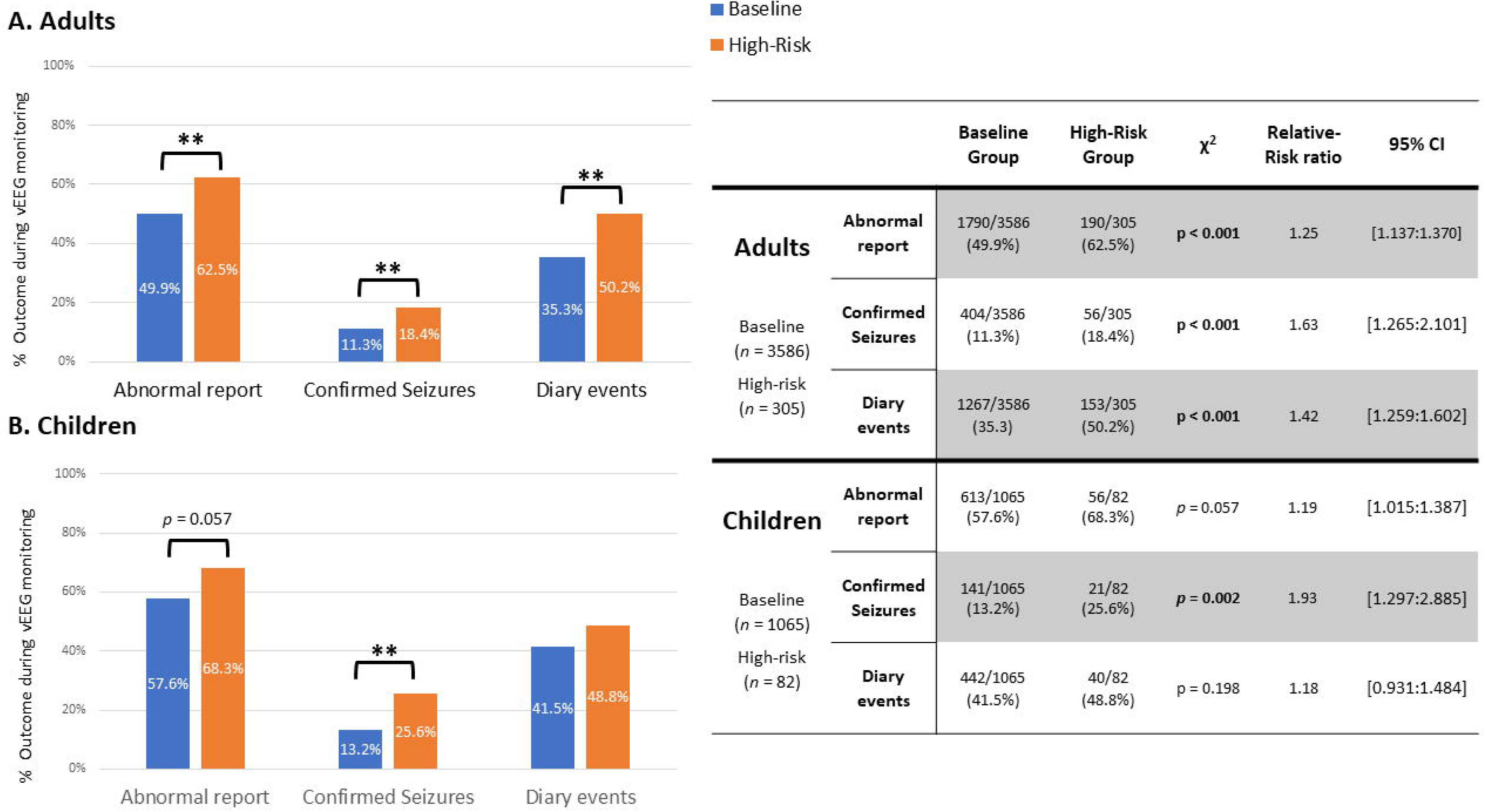
Primary analysis: Comparison of proportions for the high-risk group relative to the baseline group for both age groups (adults and children) across the three outcome measures: 1. EEG report outcome, 2. confirmed seizures, and 3. diary events. ** *p* < 0.001

#### Children

1147 children who completed ambulatory vEEG monitoring were divided into the high-risk (n=82) or baseline (n=1065) group (as presented in Figure 2B). Relative to baseline, those in high-risk were 19% more likely to present with an abnormal report (56/82:68.3% vs 613/1065:57.6%, RR=1.19, 95% CI[1.015:1.387], *p*=0.057), 93% more likely to have a confirmed seizure during vEEG monitoring (21/82:25.6% vs 141/1065:13.2%, RR=1.93, 95% CI[1.297:2.885], *p=*0.002) and 18% more likely to report a diary event during monitoring (40/82:48.8% vs 442/1065:41.5%, RR=1.18, 95% CI[0.931:1.484], *p=*0.198).

### II. Secondary Analysis (*presented in Figure 3 and Table 2*)

#### a. Gender

##### I. Females

Across a sample of 2397 females, those in high-risk relative to baseline, were 24% more likely to have an abnormal report (115/193:59.6% vs 1064/2204:48.3%, RR=1.234, 95% CI[1.090:1.397], *p=*0.003), 118% more likely to present with a confirmed seizure during vEEG monitoring (35/193:18.1% vs 183/2204:8.3%, RR=2.184, 95% CI[1.570:3.039], *p<*0.001) and 36% more likely to report a diary event during monitoring (99/193:51.3% vs 832/2204:37.7%, RR=1.359, 95% CI[1.172:1.575], *p<*0.001).

**Figure 3.**
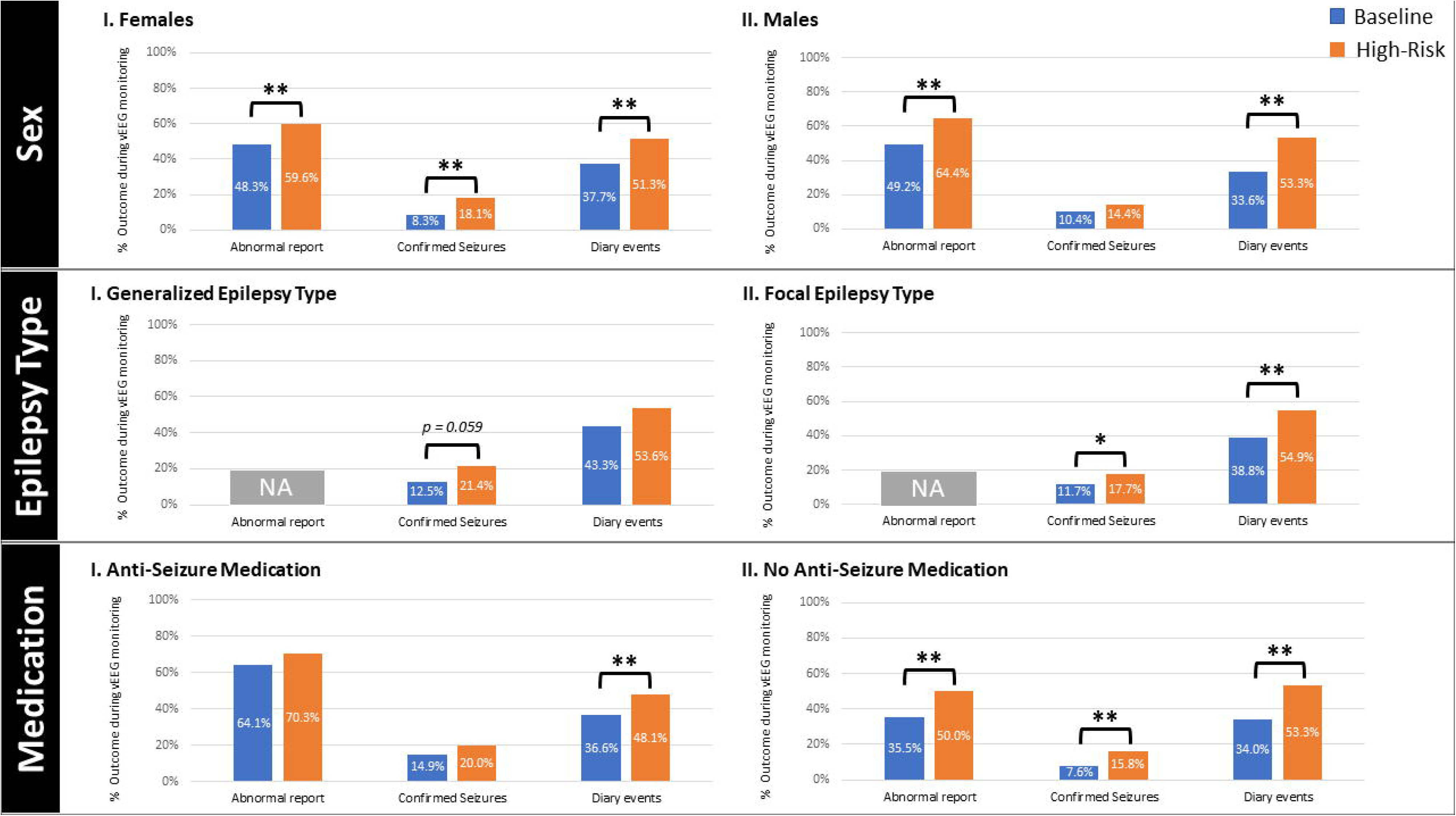
Secondary analysis: Comparisons of proportions for the high-risk group relative to the baseline group for all adults across the secondary analysis subgroups divided according to Sex (Females/Males), Epilepsy Type (Generalized/Focal), and Medication (Anti-Seizure medication/No Anti-Seizure medication reported) across the three outcome measures: 1. EEG report outcome, 2. confirmed seizures, and 3. diary events. \**p* < 0.05, ** *p* < 0.001. Note, it is not possible to compare Epilepsy Type groups with regards to an abnormal EEG report outcome given epilepsy type was divided according to information presented in the final EEG report.

**Table 2.** Secondary analysis: Presentation of data comparing proportions for the high-risk relative to the baseline group across the secondary analysis subgroups which are divided according to Sex (Females/Males), Epilepsy Type (Generalized/Focal), and Medication (Anti-Seizure medication/No Anti-Seizure medication reported) across the three outcome measures, 1. EEG report outcome, 2. Confirmed seizures, and 3. Diary events. Note, Epilepsy Type was divided according to information presented in the final EEG report, therefore, it is not possible to conduct a comparison of the Epilepsy Type groups with regards to an abnormal EEG report outcome.

| | | | Baseline | High-Risk | $\chi^2$ | Relative-Risk Ratio | 95% CI |
| --- | --- | --- | --- | --- | --- | --- | --- |
| Secondary Analysis I | Female<br>Baseline (*n* = 2204)<br>High-Risk (*n* = 193) | Abnormal report | 1064/2204 (48.3%) | 115/193 (59.6%) | *p* = 0.003 | 1.234 | [1.090:1.397] |
|  |  | Confirmed Seizures | 183/2204 (8.3%) | 35/193 (18.1%) | *p* < 0.001 | 2.184 | [1.570:3.039] |
|  |  | Diary events | 832/2204 (37.7%) | 99/193 (51.3%) | *p* < 0.001 | 1.359 | [1.172:1.575] |
|  | Male<br>Baseline (*n* = 1220)<br>High-Risk (*n* = 90) | Abnormal report | 600/1220 (49.2%) | 58/90 (64.4%) | *p* = 0.005 | 1.310 | [1.112:1.543] |
|  |  | Confirmed Seizures | 127/1220 (10.4%) | 13/90 (14.4%) | *p* = 0.232 | 1.388 | [0.817:2.355] |
|  |  | Diary events | 410/1220 (33.6%) | 48/90 (53.3%) | *p* < 0.001 | 1.587 | [1.288:1.955] |
| Secondary Analysis II | Generalized<br>Baseline (*n* = 570)<br>High-Risk (*n* = 56) | Abnormal report | NOT APPLICABLE |  |  |  |  |
|  |  | Confirmed Seizures | 71/570 (12.5%) | 12/56 (21.4%) | *p* = 0.059 | 1.720 | [0.996:2.972] |
|  |  | Diary events | 247/570 (43.3) | 30/56 (53.6%) | *p* = 0.141 | 1.236 | [0.952:1.605] |
|  | Focal<br>Baseline (*n* = 1698)<br>High-Risk (*n* = 164) | Abnormal report | NOT APPLICABLE |  |  |  |  |
|  |  | Confirmed Seizures | 198/1698 (11.7%) | 29/164 (17.7%) | *p* = 0.024 | 1.516 | [1.063:2.163] |
|  |  | Diary events | 659/1698 (38.8%) | 90/164 (54.9%) | *p* < 0.001 | 1.414 | [1.216:1.645] |
| Secondary Analysis III | Anti-Seizure Medication<br>Baseline (*n* = 1804)<br>High-Risk (*n* = 185) | Abnormal report | 1157/1804 (64.1%) | 130/185 (70.3%) | *p* = 0.096 | 1.096 | [0.992:1.211] |
|  |  | Confirmed Seizures | 268/1804 (14.9%) | 37/185 (20%) | *p* = 0.064 | 1.346 | [0.989:1.833] |
|  |  | Diary events | 661/1804 (36.6%) | 89/185 (48.1%) | *p* = 0.002 | 1.313 | [1.117:1.543] |
|  | No Anti-Seizure Medication<br>Baseline (*n* = 1782)<br>High-Risk (*n* = 120) | Abnormal report | 633/1782 (35.5%) | 60/120 (50%) | *p* = 0.001 | 1.408 | [1.165:1.701] |
|  |  | Confirmed Seizures | 136/1782 (7.6%) | 19/120 (15.8%) | *p* = 0.001 | 2.075 | [1.332:3.231] |
|  |  | Diary events | 606/1782 (34%) | 64/120 (53.3%) | *p* < 0.001 | 1.568 | [1.311:1.877] |

##### II. Males

Across a sample of 1310 males, those in high-risk as compared to baseline were 31% more likely to have an abnormal report (58/90:64.4% vs 600/1220:49.2%, RR=1.310, 95% CI[1.112:1.543], *p*=0.005), 39% more likely to present with a confirmed seizure during vEEG monitoring (13/90:14.4% vs 127/1220:10.4%, RR=1.388, 95% CI[0.817:2.355], *p*=0.232) and 59% more likely to report a diary event during monitoring (48/90:53.3% vs 410/1220:33.6%, RR=1.587, 95% CI[1.288:1.955], *p<*0.001).

#### b. Epilepsy Type

##### I. Generalized

In a sample of 626 adults where EEG diagnosis was consistent with generalized epilepsy, relative to baseline, those in high-risk were 72% more likely to present with a confirmed seizure during vEEG monitoring (12/56:21.4% vs 71/570:12.5%, RR=1.720, 95% CI[0.996:2.972], *p=*0.059) and 24% more likely to report a diary event during monitoring (30/56:53.6% vs 247/570:43.3%, RR=1.236, 95% CI[0.952:1.605], *p=*0.141).

##### II. Focal

In a sample of 1862 adults where EEG diagnosis was consistent with focal epilepsy, those in high-risk were 52% more likely to present with a confirmed seizure during vEEG monitoring (29/164:17.7% vs 198/1698:11.7%, RR=1.516, 95% CI[1.063:2.163], *p*=0.024) and 41% more likely to report a diary event during monitoring(90/164:54.9% vs 659/1698:38.8%, RR=1.414, 95% CI[1.216:1.645], *p<*0.001).

#### c. Medication

##### I. Medicated

In a sample of 1989 adults prescribed with at least one ASM, relative to baseline, those in high-risk were 10% more likely to have an abnormal report (130/185:70.3% vs 1157/1804:64.1%, RR=1.096, 95% CI[0.992:1.211], *p=*0.096), 35% more likely to present with a confirmed seizure during vEEG monitoring (37/185:20% vs 268/1804:14.9%, RR=1.346, 95% CI[ 0.989:1.833], *p=*0.064) and 31% more likely to report a diary event during monitoring (89/185:48.1% vs 661/1804:36.6%, RR=1.313, 95% CI[1.117:1.543], *p=*0.002).

##### II. Not medicated

In a sample of 1902 adults prescribed with ASM medication, relative to baseline, those in high-risk were 41% more likely to have an abnormal report (60/120:50% vs 633/1782:35.5%, RR=1.408, 95% CI[1.165:1.701], *p<*0.001), 108% more likely to present with a confirmed seizure during vEEG monitoring (19/120:15.8% vs 136/1782:7.6%, RR=2.075, 95% CI[1.332:3.231], *p<*0.001) and 57% more likely to report a diary event during monitoring (64/120:53.3% vs 606/1782:34%, RR=1.568, 95% CI[1.311:1.877], *p*<0.001).

Following this, the impact of monitoring length on outcome measures was assessed. Shorter vEEG monitoring timeframes (1-4 days), resulted in an increased likelihood of identifying a confirmed electrographic seizure when vEEG monitoring aligned with high risk. For longer monitoring timeframes (5-10 days), when monitoring coincided with high-risk, more outcomes were significant, including confirmed electrographic seizures, a conclusive abnormal report and a diary event during vEEG monitoring.

Next, the effect of seizure frequency was examined by examining those who reported “more than weekly” events and those who reported “less than weekly” events (based on information acquired from the registration form), separately. In those who reported more than weekly events, there was an increased likelihood during high-risk of observing a conclusive abnormal report, a confirmed electrographic seizure, and a diary event during vEEG monitoring. For those who reported less than weekly events, there was an increased likelihood of a conclusive abnormal EEG report during high-risk, with similar trends though not significant, for the identification of a confirmed electrographic seizure and a diary event during monitoring. These results suggest that seizure frequency does not appear to unduly influence the diagnostic outcomes identified in the current study. All data is presented in Figure 4 and Table 3.

**Figure 4.**
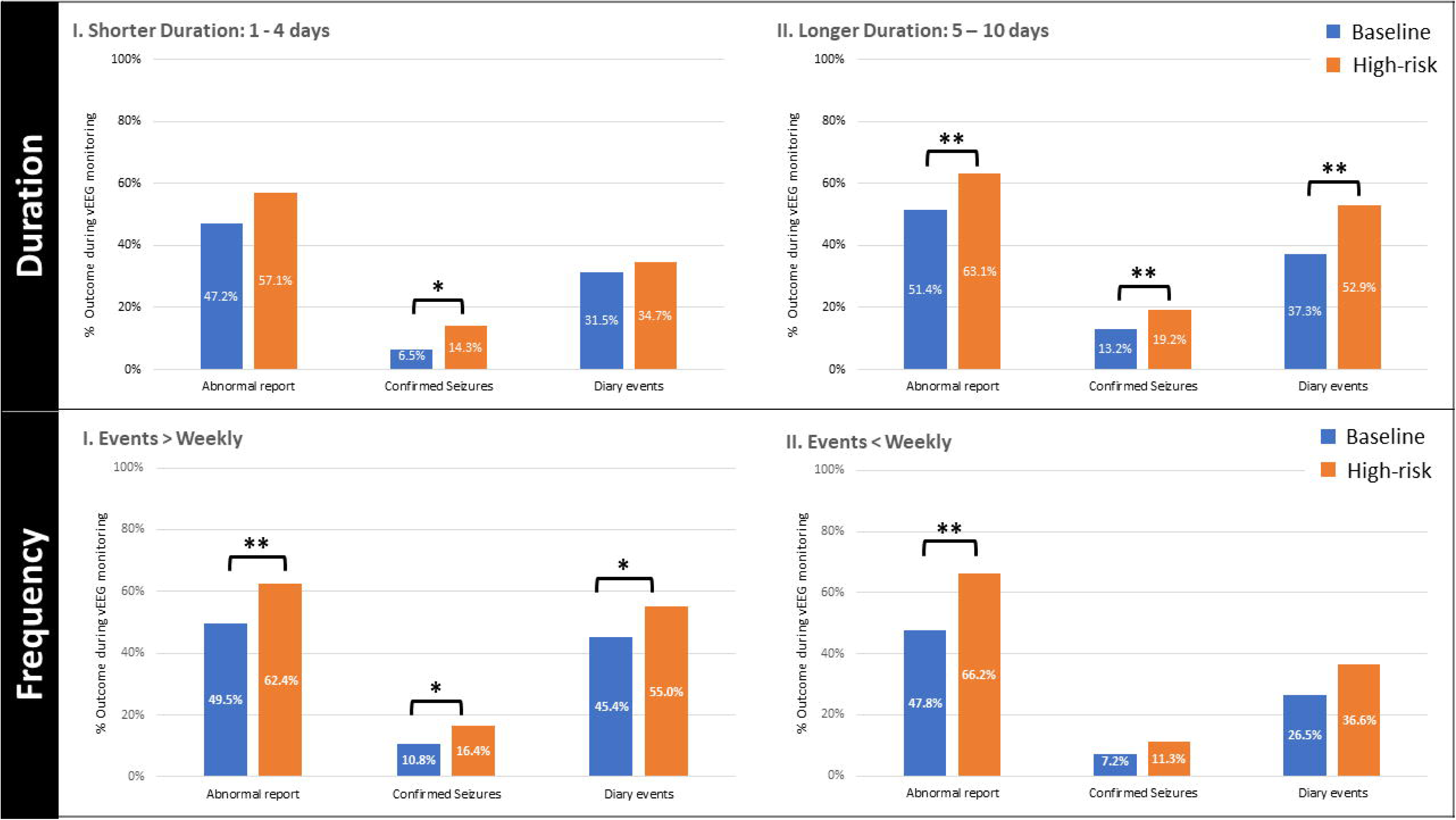
Duration and Seizure frequency: **A. Duration**: Comparison of proportions for the high-risk group relative to the comparisons baseline group for all adults divided across study length durations (Shorter durations: 1–4 days/Longer durations 5-10 days) across the three outcome measures: 1. EEG report outcome, 2. confirmed seizures, and 3. diary events, **B. Seizure frequency**: Comparison of proportions for the high-risk group relative to baseline for all adults divided across self-report measures of seizure frequency (more than weekly/less than weekly) for each risk group, separately, across the three outcome measures.

**Table 3.** Duration and self-report event frequency: Presentation of data comparing proportions for the high-risk relative to the baseline group across study length duration (Shorter durations: 1-4 days/Longer durations 5-10 days) and self-report seizure frequency (more than weekly/less than weekly) across the three outcome measures: 1. EEG report outcome, 2. confirmed seizures, and 3. diary events.

| | | | Baseline | High-Risk | $\chi^2$ | Relative-Risk Ratio | 95% CI |
| --- | --- | --- | --- | --- | --- | --- | --- |
| Duration | Shorter Duration<br>Baseline (n = 990)<br>High-Risk (n = 49) | Abnormal report | 467/990 (47.2%) | 28/49 (57.1%) | $p = 0.173$ | 1.211 | [0.942:1.557] |
| | | Confirmed Seizures | 64/990 (6.5%) | 7/49 (14.3%) | $p = 0.034$ | 2.210 | [1.070:4.566] |
| | | Diary events | 312/990 (31.5%) | 17/49 (34.7%) | $p = 0.641$ | 1.101 | [0.742:1.634] |
| | Longer Duration<br>Baseline (n = 2531)<br>High-Risk (n = 255) | Abnormal report | 1301/2531 (51.4%) | 161/255 (63.1%) | $p < 0.001$ | 1.228 | [1.241:2.113] |
| | | Confirmed Seizures | 334/2531 (13.2%) | 49/255 (19.2%) | $p = 0.008$ | 1.456 | [1.111:1.909] |
| | | Diary events | 945/2531 (37.3%) | 135/255 (52.9%) | $p < 0.001$ | 1.418 | [1.250:1.609] |
| Frequency | > Weekly<br>Baseline (n = 1763)<br>High-Risk (n = 189) | Abnormal report | 872/1763 (49.5%) | 118/189 (62.4%) | $p < 0.001$ | 1.262 | [1.119:1.424] |
| | | Confirmed Seizures | 191/1763 (10.8%) | 31/189 (16.4%) | $p = 0.022$ | 1.514 | [1.068:2.145] |
| | | Diary events | 800/1763 (45.4%) | 104/189 (55.0%) | $p = 0.011$ | 1.213 | [1.056:1.393] |
| | < Weekly<br>Baseline (n = 1689)<br>High-Risk (n = 71) | Abnormal report | 807/1689 (47.8%) | 46/71 (66.2%) | $p = 0.002$ | 1.385 | [1.165:1.648] |
| | | Confirmed Seizures | 121/1689 (7.2%) | 8/71 (11.3%) | $p = 0.194$ | 1.573 | [0.801:3.089] |
| | | Diary events | 447/1689 (26.5) | 26/71 (36.6%) | $p = 0.059$ | 1.384 | [1.009:1.898] |

## Discussion

The current study introduced pro-ictal EEG scheduling as a novel low-risk strategy for increasing the clinical yield of vEEG. Findings from this large-scale validation study provide strong support for enhanced vEEG capabilities when monitoring was aligned with estimated periods of heightened seizure risk across both adult and paediatric cohorts (Figure 2). Additionally, this innovative approach appears to have broad applicability within the adult cohort, as demonstrated by the similar results when examined across a range of clinical and demographic factors (Figure 3).

### I. Primary findings

The current study provides compelling evidence for the clinical efficacy of the pro-ictal EEG scheduling framework based on multiday seizure cycles (extracted from seizure diaries) when combined with probabilistic seizure forecasting techniques. Specifically, this is the first large-scale validation study to demonstrate that personalized estimates of heightened seizure risk coincided with an increased likelihood of capturing epileptiform activity (both interictal activity and confirmed seizures) during vEEG monitoring in an adult cohort. This was also examined within a paediatric cohort, where a challenging aspect of the diagnosis is the presence of several paroxysmal disorders with very similar features, which can lead to issues with misdiagnosis ^31^; thus, diagnosis often requires confirmation by vEEG^32^. While recent studies suggest that electronic seizure diaries ^25,27,33^ and wearable devices^15^ can be used to extrapolate multiday seizure cycles across different age groups, the current study is the first to validate the pro-ictal EEG scheduling approach in a paediatric cohort. In the paediatric cohort, when vEEG monitoring aligned with an estimated high-risk period, there was an increased likelihood of capturing a confirmed seizure during monitoring. These findings provide strong evidence across both adult and paediatric cohorts for the efficacy of pro-ictal EEG scheduling as a clinically relevant method for optimizing the diagnostic yield of vEEG.

Key clinical benefits of applying pro-ictal EEG scheduling include: 1. improving vEEG yield without the need for seizure provocation or suggestive seizure manipulation; 2. specialized hospital units and teams are not necessarily required for pro-ictal scheduling, leading to reduced stress on hospital resources and infrastructure ^8^(for instance, in the current study, pro-ictal EEG scheduling was shown to be effective at increasing capture rate when combined with ambulatory EEG) and 3. enables a naturalistic observation of spontaneous seizures without the need for active seizure induction or medication adjustments (which could potentially alter features of the recorded seizure).

Nevertheless, despite these considerable advantages, it is important to acknowledge that pro-ictal EEG scheduling does not replace existing seizure provocation techniques. Instead, it could be implemented as a preliminary step to enhance vEEG diagnostic capabilities by scheduling monitoring sessions to synchronize with projected pro-ictal periods. Moreover, for more severe cases or pre-surgical assessments, it is plausible that pro-ictal EEG scheduling models could enhance efficacy of seizure provocation techniques (for instance, combining both techniques could reduce seizure provocation severity and maintain the same seizure likelihood). Therefore, we propose that pro-ictal EEG scheduling could significantly impact the field of neurology by presenting a pragmatic and low-risk technique capable of enhancing current vEEG diagnostic capabilities.

### II. Demographic and clinical features

The current study sought to clarify whether the pro-ictal EEG scheduling model has widespread applicability across a range of demographic and clinical features (sex, epilepsy type and anti-seizure medication). With regards to sex, across numerous studies, data derived from both male and female participants assisted in the identification of multiday seizure cycles ^13,19,20,25,28^, with evidence of comparable seizure cycles observed between males and females ^13,20^. However, to the authors‘ knowledge, this is the first study to apply these seizure forecasting models separately across male and female cohorts and, in doing so, provides the first line of evidence supporting the efficacy of implementing the pro-ictal scheduling method to time monitoring to increase vEEG yield across both cohorts.

In terms of epilepsy type, early intracranial EEG seizure cycle studies were largely conducted in those with focal epilepsy ^17^, leading to increased research focus on multiday seizure cycles underlying focal epilepsy ^13,20,25^. Thus far, only a limited number of studies has examined features of multiday seizure cycles ^25^ and seizure forecasting accuracy across the generalized epilepsies ^28^. Our findings indicate that timing monitoring according to the predicted period of heightened risk state increased the yield of observing epileptiform activity during monitoring across long-term EEG studies for people with a focal epilepsy, with similar trends, though not significant, in individuals with diagnosis consistent with generalized epilepsy. While these preliminary findings are promising, further specialized studies are required to further assess the applicability of this approach across a range of epilepsy classifications.

As for medication effects, to date there is a very limited understanding of how medication influences multiday seizure cycles. However, seizure cycles appear to persist after controlling for medication effects ^25^. In the early observations of seizure cycles^34^ and preclinical research^35^, it was suggested that while anti-seizure medications (ASM) reduce the number of seizures, the temporal dynamics underlying seizure timing appear to remain consistent. The current study provides the first comparison of seizure forecasting efficacy during vEEG monitoring in those medicated with ASMs relative to those who were not medicated. In those not medicated, there was a significant increase in likelihood of capturing epileptic activity during high-risk and, while a similar trend emerged for those medicated with ASMs, the effect was not as substantial. These findings indicate that cyclic patterns underlying epileptic activity are unlikely to be driven by ASM use. Moreover, while ASMs act by dampening epileptiform activity ^36^ and reduce the incidence of seizure activity over the high-risk period, the temporal patterns underlying seizure cycles appear to remain intact. Therefore, the pro-ictal scheduling methodology appears to have broad applicability and shows potential to increase the diagnostic yield of vEEG across a range of clinical and demographic groups.

### III. Diary events

Despite inaccuracies in self-reporting of events ^37,38^, obtaining data from self-reported events is nevertheless a critical component of diagnosis (i.e., inspecting and understanding the diagnostic relevance of events). In the present study, there was an increased likelihood of a self-reported event when vEEG occurred during a timeframe of heightened seizure risk. This finding was observed regardless of sex, epilepsy type and medication status. The implications of these findings are important, as capturing such an event during vEEG monitoring will improve the diagnostic capability of clinicians to characterize whether reported events (documented in seizure diaries) are electrographic seizures or represent non-epileptic events or cardiac episodes, all of which can be verified from the vEEG monitoring session. This could lead to a more rapid identification of seizure type and potential reduction in the incidence of misdiagnosis of epilepsy ^39^. Notably however, an increased likelihood of a diary event during high risk was not observed in the paediatric cohort. It is plausible that different seizure frequencies and reporting styles between children (often carer or parent assisted) and adults could account for these differences ^40,41^. Moreover, the paediatric cohort had a considerably smaller sample size, so larger studies are warranted to further explore this finding in a paediatric population.

### IV. Limitations

A number of limitations require careful consideration. First, while seizure diaries are widely used for reporting seizure onset times, there are constraints in the reporting accuracy of documented seizures ^37,42^. Despite this, seizure diaries appear to capture sufficient data to extrapolate accurate individualized multiday cyclic-based patterns^18,20,27,28^, which can be used to generate a seizure forecast ^13,20,28^. This was confirmed by the current study, where multiday seizure patterns generated for both adult^18,28^ and paediatric^25^ cohorts with more than 10 diary events over a period of 12 months could produce seizure forecasts capable of identifying periods of increased seizure likelihood when vEEG monitoring occurred during estimated high-risk. Quite notably however, the current approach is limited to individuals who have been using a seizure diary prior to monitoring and will be less applicable to individuals undergoing vEEG monitoring following an initial seizure event.

Second, the current study provided a preliminary and indirect assessment of endogenous features of multiday seizure cycles (i.e., age-group, sex and epilepsy type). Further studies are required to confirm these pilot results in larger cohorts. More direct measures of physiological markers, such as heart rate, basal body temperature and sleep, and whether they relate to the temporal dynamics underlying multiday seizure cycles, could provide further insight into the mechanisms underlying these seizure cycles and potentially be implemented to optimize pro-ictal scheduling techniques. Finally, while the current study demonstrated the utility of applying a time-invariant seizure forecasting framework, future studies could build on these findings by exploring the efficacy of pro-ictal EEG scheduling in a naturalistic setting by conducting a *prospective* cohort study (i.e., actively scheduling monitoring timing according to high-risk periods). While it is anticipated that pro-ictal EEG scheduling models will continue to evolve, findings from the current study are extremely promising and present a low-risk approach which could be implemented in coordination with compatible health care systems to potentially increase the diagnostic yield of vEEG.

### VI. Conclusion

In summary, we have introduced the concept of pro-ictal EEG scheduling as a promising, low-risk strategy to enhance the diagnostic capabilities of vEEG by aligning monitoring with anticipated periods of heightened seizure risk. Improvements in the diagnostic yield of vEEG, even marginally, could have a profound effect on reducing the incidence of misdiagnosis, diagnostic delays and inconclusive reports. This study provides large-scale validation and compelling evidence for the effectiveness of pro-ictal EEG scheduling across adult and paediatric populations. This innovative approach may significantly impact neurological clinical practice by providing a robust and easily implementable method for optimizing the diagnostic capabilities of vEEG monitoring.

## Data Availability

All data produced in the present study are available upon reasonable request to the authors.

